# Effect of the foot-strike pattern on temporal characteristics of sagittal plane knee kinetics and kinematics during the early phase of cutting movements

**DOI:** 10.1101/2021.10.20.21265090

**Authors:** Yuki Uno, Issei Ogasawara, Shoji Konda, Kaito Wakabayashi, Miyakawa Motoi, Megumi Nambo, Kaho Umegaki, Haotian Cheng, Ken Hashizume, Ken Nakata

**Author notes:** **Corresponding author:** Ken Nakata and Issei Ogasawara, Department of Health and Sport Sciences, Graduate School of Medicine, Osaka University, 1-17, Machikaneyama-cho, Toyonaka city, Osaka, 560-0043, Japan.

## Abstract

Anterior cruciate ligament (ACL) injury occurs soon after foot-strike. Cutting with a shallow flexed knee is considered a risk factor for ACL injury; however, how foot-strike patterns (forefoot strike [FFS] vs. rearfoot strike [RFS]) affect sagittal plane knee kinetics and kinematics after a foot-strike, is unknown. This study aimed to investigate the effect of foot-strike patterns on the temporal characteristics in sagittal plane knee kinetics and kinematics during cutting. Twenty-three males performed 45° cutting under RFS and FFS conditions. The marker position data on the lower limb, and the ground reaction force (GRF) data were collected and time-normalized (0%–100%) during the stance phase. The knee flexion angle, shank and GRF vector inclination angle relative to the global vertical axis, knee flexion/extension moment, and anterior/posterior component of GRF relative to the shank segment were calculated and compared between foot-strike patterns using statistical parametric mapping paired *t*-test (p<0.0071). The knee flexion angle was smaller in the RFS than in the FFS in the initial 40% of the stance phase. In the RFS condition, the GRF vector was directed anteriorly to the shank segment, and the knee extension moment was produced by GRF in 0%– 7% of the stance phase; these results were not observed in the FFS condition. These results suggest that compared to FFS, RFS induces a shallow flexed knee with an anterior-directed GRF component in the early stance phase, and might potentially provoke a risk of ACL injury.

**Highlights:**

- Sagittal plane knee mechanics differed between foot-strike patterns.
- Shallower knee flexion occurred in rearfoot strike than in forefoot strike in cutting.
- Ground reaction force vector directed anteriorly to shank axis in rearfoot strike.
- Larger Knee extension moment occurred after initial contact in rearfoot strike.
- Rearfoot strike had a potentially higher risk for ACL injury than forefoot strike.

## Introduction

Anterior cruciate ligament (ACL) injury is one of the most common types of knee trauma, occurring at a rate of 68.6 per 100,000 person per year in the United States (Sanders et al., 2016). In terms of age and sex, the incidence is highest in males 19–25 years old (241.0 per 100,000 person per year). Sixty-five percent of ACL injuries are related to sports activities (Gianotti et al., 2009). Once athletes suffer from ACL injury, reconstruction surgery and long-term rehabilitation are required to regain and establish the structural and physiological function of the ACL (Adern et al., 2011). Additionally, ACL injury increases the risk of re-rupture after returning to competition (Paterno et al., 2014; Wiggins et al., 2016), subsequently resulting in a lower quality of life, including the early onset of knee osteoarthritis (Lohmander et al., 2004; von Porat et al., 2004). Although researchers and sports-persons have been challenged with preventive interventions (Padua et al., 2018), a large number of ACL injuries still occur (Bram et al., 2021; Agel et al., 2016). To establish more efficient prevention methods, we need to understand the movements associated with risky knee kinematics, kinetics, and subsequent ACL stress in sporting situations.

ACL injuries frequently occur in ball games such as basketball, which involve repetitive landings and cuttings (Griffin et al., 2000), and an early deceleration phase immediately after initial foot contact (IC)—approximately 50 ms from IC—is the most provocative time frame for ACL injury (Krosshaug et al., 2007). Approximately 70%–80% of ACL injuries in sports occur in a noncontact or indirect contact manner (Boden et al., 2000; Renstrom et al., 2008; Waldén et al., 2015). Collectively, these characteristics allow us to predict that the rapidly developed ground reaction force (GRF) acting on the braking limb is a significant mechanical source for promoting a risky knee loading status (Shimokochi and Shultz, 2008). A small knee flexion angle has been frequently observed during ACL injury (Boden et al., 2010; Della Villa et al., 2020; Montgomery et al., 2016), and it is known that the externally applied knee forces and moments increases the ACL tension particularly at the shallow flexed knee position (Markolf et al., 1995; Sakane et al., 1997). These findings suggest that the shallow flexed knee position under increased GRF after IC is the highest risk situation for a non-contact ACL injury. Therefore, elucidating the technical factors that induce a shallow flexed knee during the early deceleration phase is essential to an injury prevention regimen.

The foot-strike pattern is a potential technical factor that influences knee flexion angle and the sagittal plane knee mechanics of the braking limb, and may be associated with the risk of ACL injury (Shimokochi et al., 2016). The rearfoot strike (RFS) is considered a high-risk technique for ACL injury because ACL injury is more frequently observed in a RFS than a forefoot strike (FFS) (Della Villa et al., 2020; Montgomery et al., 2016). Lab-controlled studies suggested that RFS more frequently produces combined knee valgus and tibial internal rotation moment compared to FFS (Ogasawara et al., 2020) and the peak knee valgus angle and valgus moments were significantly greater in RFS than in FFS during cuttings (Cortes et al., 2012; David et al., 2017). Focusing on the knee sagittal plane, Cortes et al. (2012) found that the knee flexion angle was smaller in RFS than FFS, at the moment of IC. Although it is known that ACL injury three-dimensionally develops within a certain time after foot strike, (Koga et al., 2010; Quatman and Hewett, 2009; Shimokochi and Shultz, 2008), due to its anatomical structure, the injury sequence may be largely influenced by sagittal mechanics such as anterior tibial shear force with a shallow knee flexed position (Fleming et al., 2001). However, to the best of our knowledge, only a few studies have investigated the effect of foot-strike pattern on the temporal characteristics of the sagittal plane knee kinetics and kinematics, as well as the potential risk of ACL injury in the early (deceleration) phase of cutting maneuvers.

The purpose of this study was to examine the effect of foot-strike pattern (RFS vs. FFS) on the temporal characteristics of sagittal plane knee kinematics and kinetics during the early phase of cutting movements. Considering the risk of RFS, we hypothesized that i) the knee flexion angle remains smaller in RFS than FFS after IC, ii) the shank segment of the stance limb inclines more posterior in RFS than FFS, iii) the line of action of the GRF vector is directed the anterior to the shank segment of the stance limb in RFS but not in FFS, and iv) the knee resultant extension moment applies in RFS but not in FFS, during the early stance phase.

## Methods

### Participants

Twenty-three healthy male athletes (mean height, 172.9 [SD 4.9] cm; mean weight, 67.8 [SD 6.4] kg; mean age, 20.2 [SD 1.3] years old) participated. A priori power analysis was performed to determine the sample size required to achieve 80% power at a statistical significance criterion of 0.05, with a large effect size of Cohen’s d=0.8 using G*Power 3.1.9.3 (Faul et al., 2007); it provided N=23 as a sample size for each foot-strike condition. All participants were recreational athletes who had more than four years of experience in handball or basketball and were familiar with cutting maneuvers. None of the participants had a previous history of severe lower-limb musculoskeletal injuries (such as ACL tears) and no minor lower-limb trauma within 6 months. This study was approved by the Ethics Committee of Osaka University Hospital (18082), and written informed consent was obtained from all participants.

### Procedure

All participants wore a black spandex shirt, pants, and the same model of shoes (THH544-001, ASICS, Japan). Seventeen reflective markers (Table 1) were placed on the participant’s anatomical landmarks using double-sided tape (Fig. 1). One researcher carefully assessed the accuracy of marker placement for inter-participant consistency. Participants were asked to perform a 2-m approach run on a wooden platform, followed by a 45° direction change on the force plate with their dominant leg (Cortes et al., 2012). Leg dominance was determined based on the participant’s preference for single-leg cutting; 11 participants selected their right leg as their preferred leg and the others selected their left leg.

**Table 1.** Location of the reflective markers.

| Position name | Definition of marker position |
| --- | --- |
| Toe | Most anterior point of toe part of shoe |
| Medial toe | Most prominent point of the first metatarsophalangeal joint, over the shoe |
| Lateral toe | Most prominent point of the fifth metatarsophalangeal joint, over the shoe |
| Medial ankle | Most prominent point of the medial malleolus |
| Lateral ankle | Most prominent point of the lateral malleolus |
| Heel | Most posterior point of the shoe midsole |
| Medial knee | Most prominent point of the medial femoral condyle |
| Lateral knee | Most prominent point of the lateral femoral condyle |
| Tibial tuberosity | Most prominent point of the anterior aspect of the tibial tuberosity |
| Great trochanter | Most prominent point of the great trochanters (Both side) |
| Anterior thigh | Middle point of the anterior aspect of the thigh |
| ASIS | Most prominent point of the anterior superior iliac spines |
| PSIS | Most prominent point of the posterior superior iliac spines |

**Figure 1.**
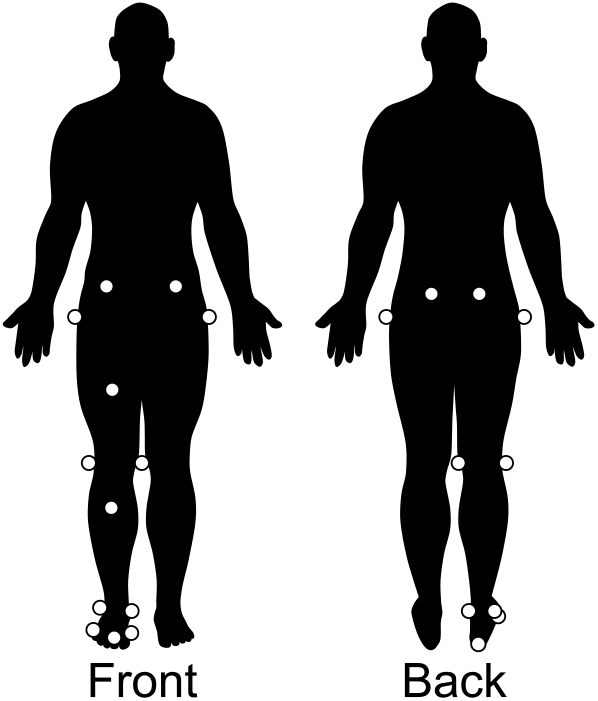
Location of the reflective marker positions. *Note that this marker set was for the right leg user.

The cutting task was performed under two different foot-strike conditions (RFS and FFS). In the RFS condition, the participants were instructed to strike their heel first with the force plate and then subsequently shift their foot-floor contact point to their forefoot for propulsion (Fig. 2). In the FFS condition, the participants were asked to touch the ball of their foot with the force plate throughout the stance phase and not to place the foot-floor contact point behind the shank axis (Fig. 2). The cutting destination of 45° from the approach-line toward the opposite of the stance leg was shown with the tape on the platform. The approach speed was set within 2.0–3.0 m/s range. Although we acknowledged that our approach speed was slightly less than those of previous similar studies (Cortes et al., 2012; McLean et al., 2005), since the RFS has been reported to promote a risky knee-loading pattern (Ogasawara et al., 2020), we controlled the approach speed to reduce the magnitude of GRF for participants’ safety. The average running speed 0.6 m prior to the force plate was monitored online using the phototube sensors with a custom LabVIEW script (LabVIEW 2016, National Instruments, Austin, TX, USA). After five minutes of warm-up, the participants familiarized themselves with the task requirements which were as follows: the correct foot-strike condition on the force plate, the appropriate approach speed (2.0–3.0 m/s), and the angle of the direction change. The order of the two foot-strike conditions was randomized for each participant, and data from seven successful trials were collected for each foot-strike condition. The task quality was judged by two experimenters, and if no consensus could be reached, the trial was discarded and an additional trial was requested. A two-minute rest occurred between foot-strike conditions and an approximately 15-second rest-period was provided between trials to reduce the effect of fatigue. The three-dimensional (3D) position of each reflective marker was captured with 11 optical cameras (OptiTrack Prime 17W, NaturalPoint, Inc., Corvallis, OR, USA) with a 360 Hz sampling frequency. The GRF was measured with a force plate (BP600400, Advanced Mechanical Technology, Inc., Watertown, MA, USA) and sampled using PowerLab (ADInstruments, New South Wales, Australia) at a 2k Hz sampling frequency. Both signals were synchronized using eSync2 (NaturalPoint, Inc., Corvallis, OR, USA).

**Figure 2.**
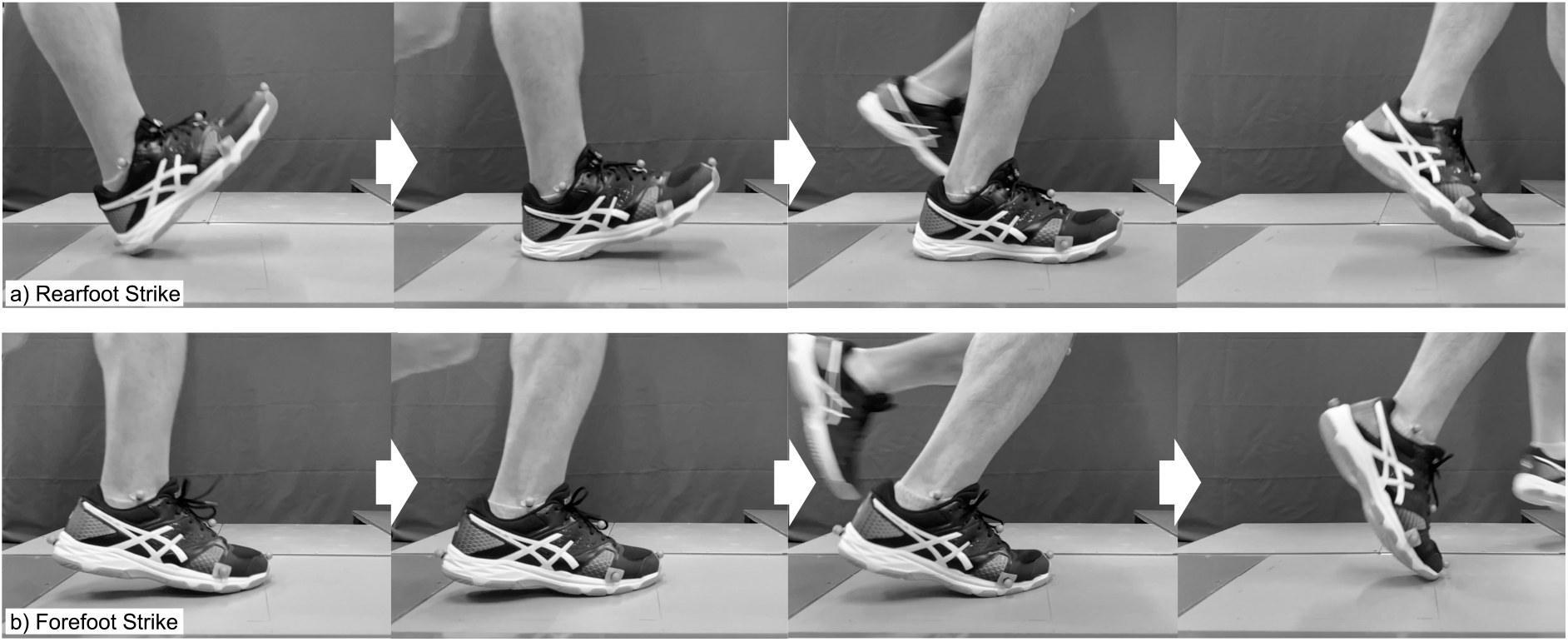
Foot-strike conditions. a) Rearfoot strike condition: the participants were instructed to strike their heel first with the force plate and subsequently shift their foot-floor contact point to their forefoot, for propulsion. b) Forefoot strike condition: the participants were requested to touch the ball of their foot with the force plate throughout the stance phase and not to place the foot-floor contact point behind the shank axis.

### Data processing

The marker and force-plate data were smoothed with a second order Butterworth digital filter (low-pass, zero-lag, cutoff frequency: 10 Hz, 50 Hz, respectively). The optimal cut-off frequencies for the marker and the force-plate data were finally determined by the biomechanics experts (IO) in terms of the balance of noise removal and signal preservation, especially around the foot-contact phase. The force-plate data were interpolated using a spline function to obtain 360 Hz resampled data. The marker and force-plate data for the stance phase (vertical GRF > 10 N) were extracted and time-normalized to 101 points (0%–100%). Using the marker data, a 3D kinematic model comprising three segments (foot, shank, and thigh) and two joints (ankle and knee) was constructed. The local coordinate systems (LCS) were defined on the shank and thigh segments (Fig. 3), and the segmental mass, position of mass center, and inertia moment for each segment were referenced to the Japanese athlete data provided by Ae et al. (1990). The kinematic variables of interest were as follows: i) knee flexion/extension angle, ii) shank inclination angle relative to the global vertical axis, iii) GRF inclination angle relative to the global vertical axis, and iv) center of pressure (CoP) anterior/posterior position relative to the shank proximal/distal axis. The kinetic variables assessed were as follows: i) knee resultant flexion/extension moment, ii) knee flexion/extension moment due to GRF, and iii) anterior/posterior component of the GRF vector relative to the shank proximal/distal axis (Fig. 3, Supplementary material). The numerical calculations were performed using custom scripts of Scilab branch-6.1.

**Figure 3.**
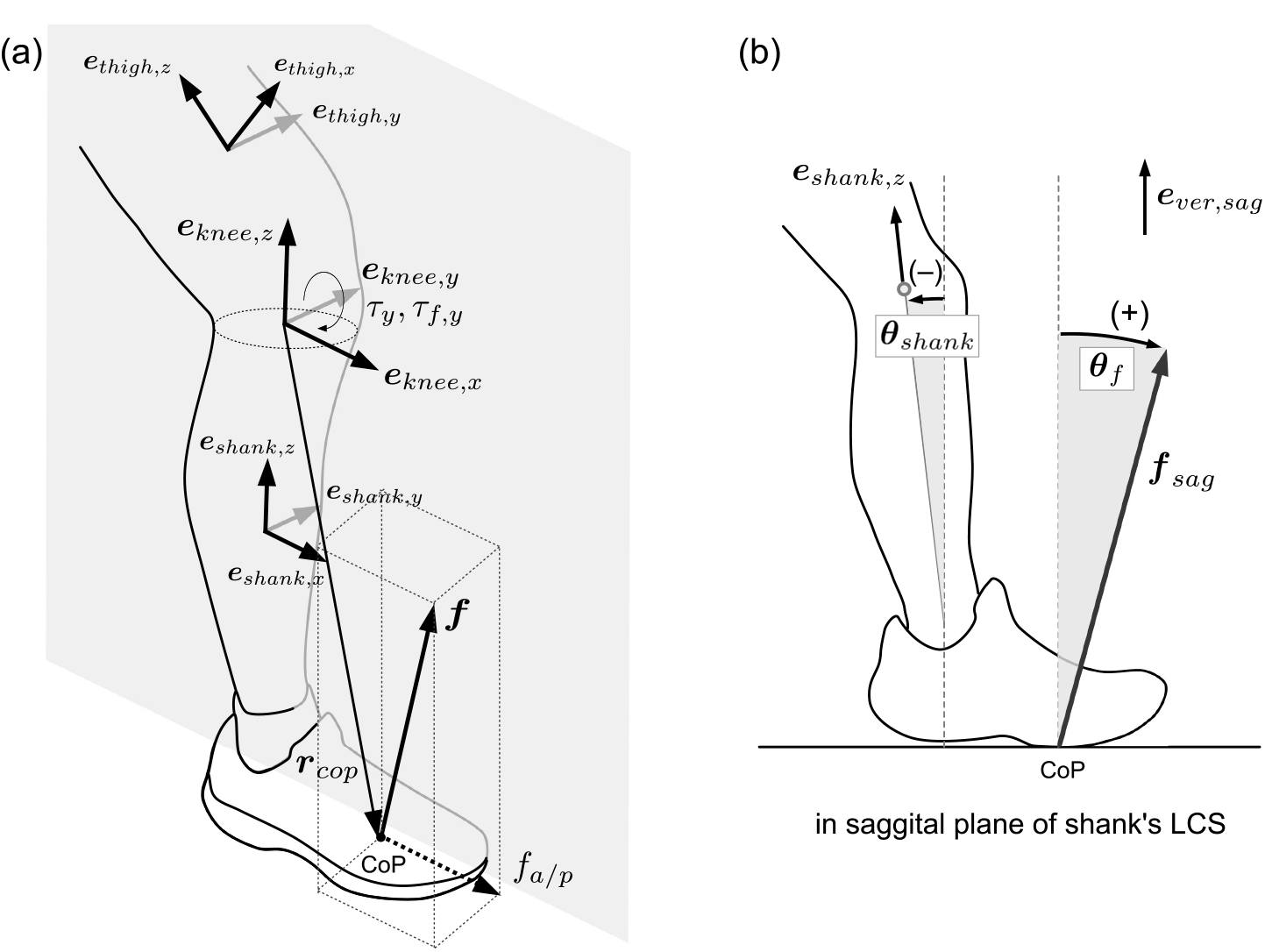
Definition of local coordinate system, knee joint coordinate system, and variables. The local coordinate systems (LCS) of the shank and thigh segments were defined by three unit-base vectors (***e***_*j,x*_, ***e***_*j,y*_, ***e***_*j,z*_, *j* = *shank, thigh*). The unit-base vector ***e***_*j,x*_ is the anterior/posterior axis pointing anterior, ***e***_*j,y*_was the medial/lateral axis pointing to the left side of the participant, and ***e***_*j,z*_was the proximal/distal axis pointing proximally. The origins of the LCS were fixed at the proximal tips of each segment. The knee joint coordinate system (KJCS) was defined by three unit vectors (***e***_*knee,x*_, ***e***_*knee,y*_, ***e***_*knee,z*_), where ***e***_*knee,y*_ is ***e***_*thigh,y*_, ***e***_*knee,z*_ is ***e***_*shank,z*_, and ***e***_*knee,x*_ is the cross product of ***e***_*knee,y*_ and ***e***_*knee,x*_ pointing anterior. The mediolateral axis of the KJCS ***e***_*knee,y*_ was defined as the knee flexion/extension axis. ***r***_*cop*_ is the position vector from KJC to the CoP. *τ*_*y*_ is the knee resultant flexion/extension moment expressed in the shank’s LCS. *τ*_2,*y*_ is the knee flexion/extension moment due to the ground reaction force (GRF) expressed in the LCS of the shank. *f*_*a/p*_ is the anterior/posterior component of the GRF vector relative to the shank proximal/distal axis. *θ*_*shank*_ and *θ*_*f*_ are the shank inclination angle and GRF inclination angle relative to the global vertical axis, respectively. See supplementary material for the detailed variable calculations.

### Statistical analysis

To confirm whether the approach speeds differed between foot-strike conditions, the average speed of the midpoint of the two ASIS markers during the 100 ms prior to the IC was compared between the conditions (paired *t*-test, p<0.01). The stance phase duration was also compared between the conditions (paired *t*-test, p<0.01). The time-series data of the knee flexion/extension angle, knee resultant flexion/extension moment, knee flexion/extension moment due to GRF, anterior/posterior component of GRF vector, and the anterior/posterior CoP position were calculated as ensemble average and standard deviation, and compared between foot-strike conditions using the one-dimensional statistical parametric mapping (SPM) two-tailed paired *t*-test (Pataky et al., 2013). The alpha level was adjusted to 0.0071 for a seven-variable comparison (Robinson et al., 2014). SPM analyses were conducted using the open-source code (spm1d) in Python 3.8.1.

## Results

Approach speed and stance phase duration

The approach speed did not differ significantly between foot-strike conditions (RFS: 2.96 [SD 0.26] m/s, FFS: 3.01 [SD 0.24] m/s, p=0.253). The stance phase duration was significantly longer in RFS than in FFS (RFS: 302.4 [SD 31.2] ms, FFS: 278.8 [SD 36.1] ms, p<0.001).

### Kinematic variables

In the RFS condition, a significantly smaller knee flexion angle was observed in 0%–39% of the stance phase, compared to the FFS condition (p<0.001, Fig. 4a). For both foot-strike conditions, the shank inclined posteriorly relative to the global vertical axis at the IC and gradually leaned to the anterior. A more posteriorly inclined shank angle was observed in RFS than in FFS at 0%–77 % of stance phase (p<0.001, Fig. 4f). For the GRF inclination angle, there was no significant difference in IC between conditions; however, the GRF vector in the RFS anteriorly inclined, whereas it tended to incline posteriorly in FFS (Fig. 4g), showing two significantly different phases (3%–7% and 53%–76%, p<0.001). In the RFS condition, the CoP was located posterior to the shank axis and then moved anteriorly, whereas in FFS, the CoP was located anterior to the shank axis throughout the stance phase. The CoP position in the RFS was significantly posterior to that in the FFS (0%–65%, p<0.001, Fig. 4 d).

**Figure 4.**
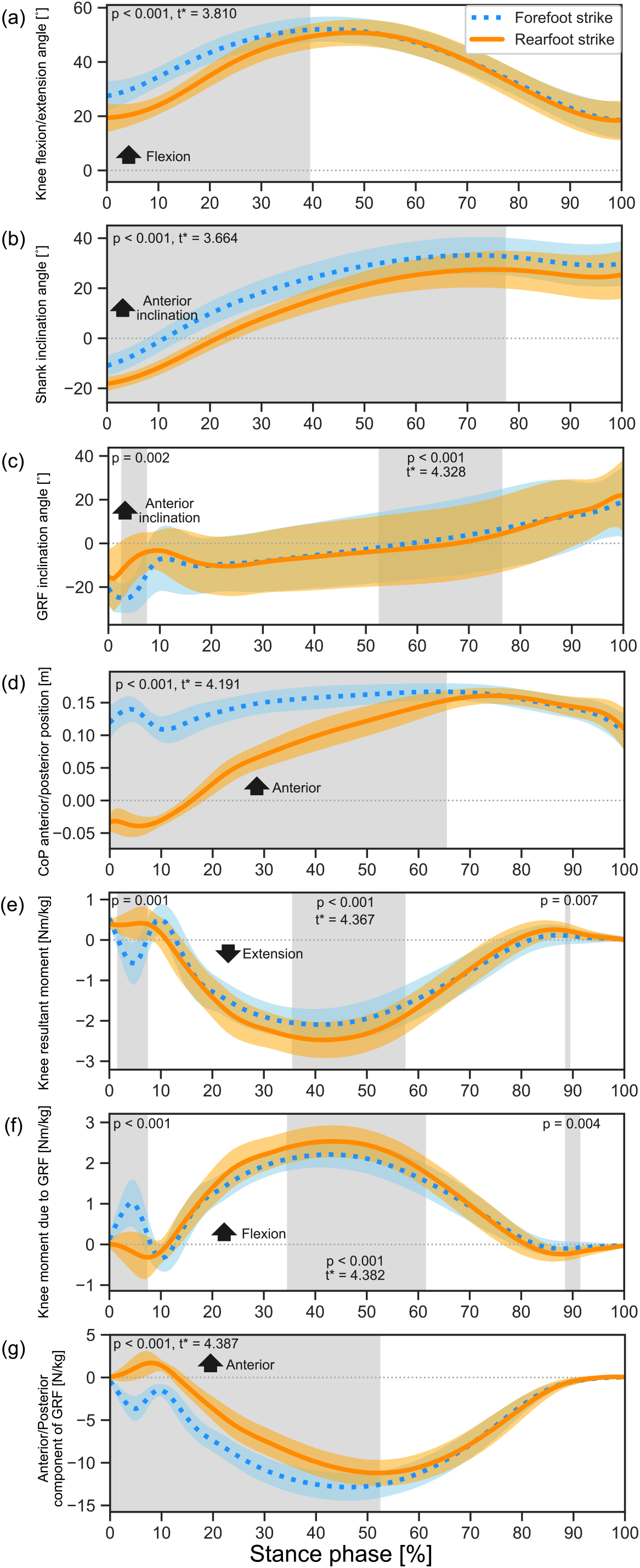
Comparison of kinematic and kinetic time-series variables between the foot-strike conditions. Time-series data of knee flexion/extension angle, shank inclination angle, GRF inclination angle, CoP anterior/posterior position, knee resultant flexion/extension moment, knee moment due to ground reaction force (GRF), and anterior/posterior component of GRF. The orange solid line and orange shadow indicate the ensemble average and standard deviation of the rear foot strike condition, and the blue dotted line and blue shadow indicate the forefoot strike condition. The gray shadow indicates the duration at which the statistical parametric mapping test revealed a significant difference between foot-strike conditions with an alpha level less than 0.0071. For each duration that showed a significant difference, the p-value and critical threshold (t*) were denoted.

### Kinetic variables

The knee resultant moment and moment due to the GRF showed similar temporal change patterns among each foot-strike pattern. With RFS, the knee resultant moment was extension direction after IC, then changed to flexion direction; with FFS, after IC, the knee resultant moment showed flexion, extension, and flexion direction in order. In the SPM results, there were significant differences in the following three duration periods: 2%–7% (p<0.001), 35%– 57% (p<0.001), and 89% (p=0.007) (Fig. 4e). The knee moment due to GRF showed almost the same statistical results (0%–7% [p<0.001], 35%–61% [p<0.001], and 89%–91% [p=0.004], Fig. 4f). The GRF vector was oppositely directed between foot-strike conditions after IC. With RFS, GRF acted in the anterior direction and then changed to a posterior direction in the shank’s LCS; with FFS, GRF acted in the posterior direction in the shank’s LCS (0%–52%, p<0.001, Fig. 4g).

## Discussion

Our results showed that RFS produced a significantly smaller knee flexion angle and more posteriorly inclined shank orientation compared with FFS during the early stance phase. The line of action of the GRF vector relative to the shank segment in the knee sagittal plane was opposite between foot-strike patterns: the GRF vector was directed anteriorly to the shank in RFS, whereas it was directed posteriorly in FFS just after IC. This kinematic relationship between the shank segment and the GRF vector in the knee sagittal plane resulted in a characteristic difference in the direction of the knee resultant moment at 2%–7 % of the stance phase. These results supported all of our hypotheses. The participants successfully controlled foot-floor contact point as the initial CoP positions for FFS and RFS were anterior and posterior to the shank segment, respectively (Fig. 4d). The temporal characteristics of the anterior/posterior CoP positions and the knee flexion angles for both foot-strike patterns were consistent with the previous cutting studies that investigated foot-strike patterns (Cortes et al., 2012; Ogasawara et al., 2020), suggesting that the participants of this study reproduced the equivalent quality of cutting and foot-strikes as that in previous studies.

The interesting point in the temporal characteristics of the knee resultant moment was that the FFS exhibited a characteristic rapid increase in the knee flexion moment within the first 10% of the stance, while the RFS showed a statistically significant and constant knee extension moment within the same time frame (Fig. 4e). The reason for the different temporal patterns in the knee resultant moment between the two foot-strike patterns is that the shank segment of the stance limb received a rapidly increased posterior-directed GRF vector component in the first 10% of stance in the FFS, whereas in the RFS condition, the anterior-directed constant GRF vector component was applied (Fig. 4g, 5). These two opposing GRF vector components produced an opposing moment due to the GRF vector around the knee (Fig. 4f), and to maintain the sagittal plane knee position against the externally applied moment due to the GRF vector, the knee resultant moment produced a counteracting direction of the moment during the early stance phase of cutting (Fig. 4e). In addition to the difference in the GRF vector orientations, the different shank orientations for FFS and RFS also explain the opposing knee resultant moment directions between the foot-strike conditions. Both foot-strike conditions showed a posteriorly inclined shank orientation relative to the global vertical axis at IC, but the FFS produced a more anteriorly inclined shank orientation than the RFS (Fig. 4b). Collectively, the RFS and FFS are foot-strike patterns that not only differentiate the foot-floor contact point, but also affect the kinematic relationship between the GRF vector and the shank segment in the knee sagittal plane at the IC, resulting in the opposite direction of the knee moment in the sagittal plane.

**Figure 5.**
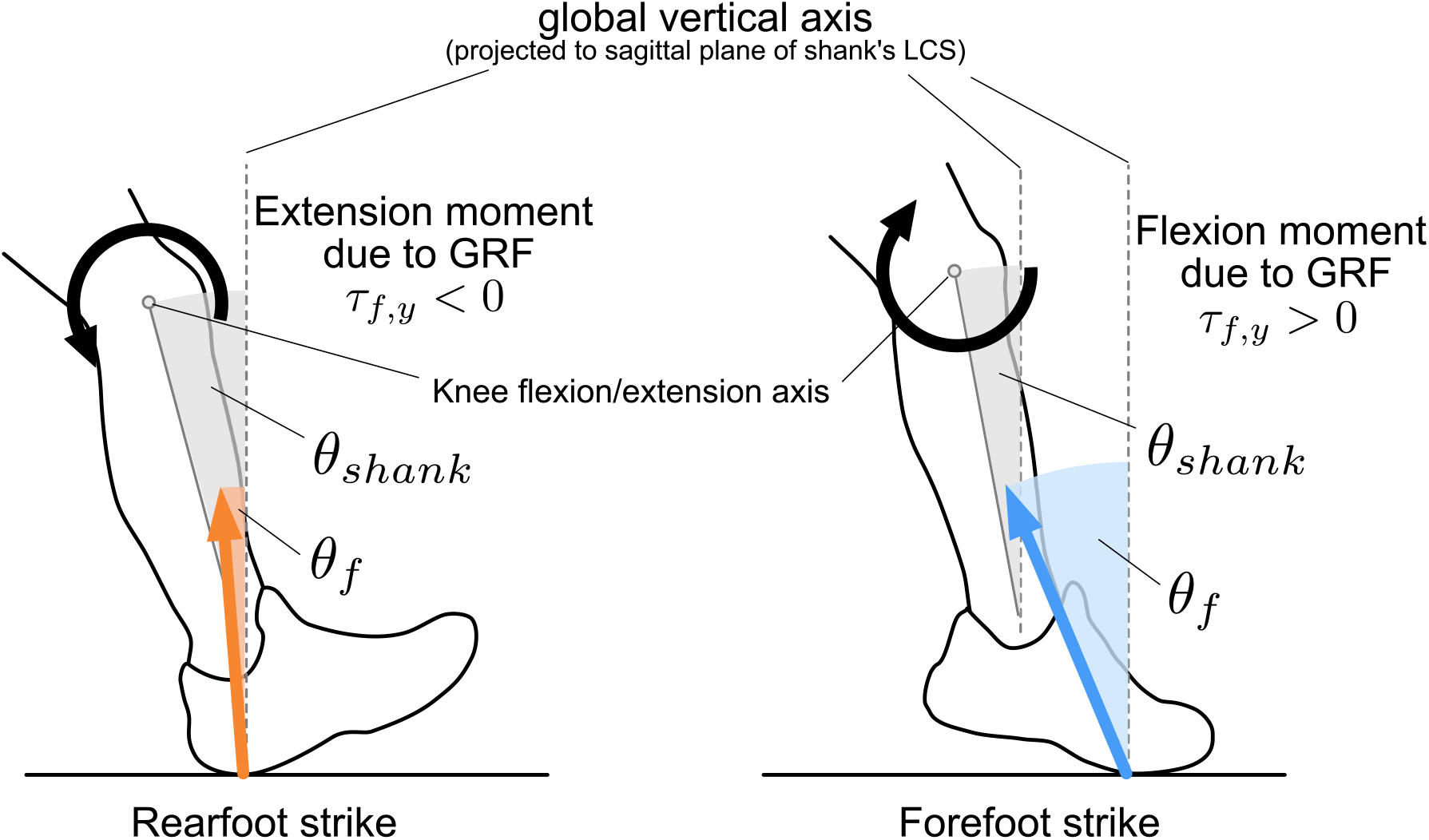
The kinematic relationship among the global vertical axis, the ground reaction force (GRF) vector, the shank segment, and the resulting direction of the moment due to GRF acting at the proximal end of the shank segment in the first 10% of stance phase. In the rearfoot strike (RFS) condition (left), the shank inclined more posteriorly and the GRF vector was directed more upright (orange arrow); then, the GRF vector produced the knee extension moment. In contrast, in the forefoot strike condition (right), the shank segment’s posterior inclination was smaller than that in the RFS, and the GRF vector was directed posterior to the shank segment (blue arrow); therefore, the GRF vector produced the knee flexion moment.

Herein, the knee flexion angle was significantly smaller in the RFS than in the FFS until 40% of the stance phase, and the angle for RFS was less than 30° until approximately 15% of the stance phase. The smaller knee flexion angle observed in the early stance phase in the RFS might potentially add to the vulnerability of the ACL, compared to the FFS, in cases of abnormal knee external stress applied to the knee during cuttings. Reportedly, the externally applied tibial internal rotation moment combined with the tibial anterior drawer force, specifically increased the ACL *in situ* force at less than 30° of knee flexion angle (Markolf et al., 1995; Sakane et al., 1997). Moreover, the patellar tendon force is converted into a larger anterior drawer force when the knee flexion angle is small (Englander et al., 2019). Herein, the first 15% of the stance phase was approximately equivalent to 40 ms on a real-time scale, and this time frame is the most provocative period for ACL injury (Krosshaug et al., 2007). Studies have reported that the peak knee valgus moment and the peak internal rotation moment occurred in the first 15% of stance phase in cuttings (McLean et al., 2005; Ogasawara et al., 2020; Sigward and Powers, 2007). *In vitro* simulation studies have reported that the peak ACL strain occurs within 40 ms after the initiation of the landing impact force (Shin et al., 2007; Withrow et al., 2006). Collectively, the RFS may be a potential foot-strike technique that makes the ACL more prone to external knee stress over producing a shallow flexed knee during the early contact phase in cutting, compared to the FFS.

The anterior/posterior component of the GRF vector relative to the shank segment was oppositely directed between RFS and FFS in the early stance phase (Fig. 4g), suggesting that the tibial anterior/posterior translational mechanics may be also influenced by different foot-strike patterns. We believe that the posterior-directed GRF vector observed in the FFS did not contribute to an increase in ACL force because this direction of force component pushes the tibia posteriorly during the braking phase, while the upper body parts, including the femur, still move forward owing to conservation of its momentum. This instantaneous mechanical discrepancy between the tibia and femur in the sagittal plane of the knee might slack the ACL. Previous simulation studies have shown that the posterior-directed GRF component reduces the ACL force during simulated double-leg landing, and its effect on ACL force was greater than that of the quadricep’s anterior drawer force (Pflum et al., 2004). Furthermore, the ACL strain was minimized when the posterior-directed GRF was applied compared to when the posterior-directed GRF was not applied in the study mimicking landing motion using cadaveric knees (Shin et al., 2007). Based on previous reports that support the idea that posterior-directed GRF relieves ACL stress, combined with our findings, indicating that the FFS produced instantaneous posterior-directed GRF at a provocative period for ACL injury, we recommend the acquisition of a FFS technique as an ACL protective foot-landing skill in accordance with previous studies (Ogasawara et al., 2020; Shimokochi et al., 2016).

This study has some limitations. Since the approach speed of 2–3 m/s was slower than that in previous studies (Cortes et al., 2012), caution should be taken when generalizing the magnitude of GRFs and knee moments. Although the posterior component of GRF may become larger with a faster approach speed, in a previous study that executed cutting with greater approach speed (4.5–5.5 m/s), GRF acted anteriorly relative to the shank during the early phase of the RFS cutting (McLean et al., 2004a). Even with slower approach speeds, the quality of the cutting movement in this study is warranted, and our suggestion about the relative direction of the GRF and moment would not change. Since this study investigated male athletes only, the results of this study should be applied with caution to female athletes. Previous video studies targeted male athletes and reported that ACL injuries occurred frequently with RFS (Della Villa et al., 2020; Montgomery et al., 2016). We investigated the effect of foot-strike patterns among male athletes and used a single-sex design to eliminate the effect of sex differences because female athletes are reported to have a smaller knee flexion angle during the cutting movement compared with their male counterparts (McLean et al., 2004b; Sigward and Powers, 2006).

In conclusion, the effect of foot-strike patterns was demonstrated on the sagittal plane knee kinematics and kinetics during the cutting movement. Compared with FFS, RFS is associated with the anterior-directed GRF relative to the shank segment, resulting in the extension moment due to GRF, while knee flexion angle remained smaller after IC. These kinematic and kinetic characteristics of RFS potentially place the ACL more prone to mechanical stress compared to FFS, in the case of application of abnormal external knee stress.

## Data Availability

All data produced in the present study are available upon reasonable request to the authors

