## Supplementary Material for "Effect of the foot-strike pattern on temporal characteristics of sagittal plane knee kinetics and kinematics during the early phase of cutting movements"

Definition of kinematic and kinetic variables

The knee flexion/extension angle was calculated using Grood and Suntay's method (Grood and Suntay, 1983). The shank inclination angle $\theta_{shank}$ was defined as an angle between $\boldsymbol{e}_{shank,z}$ and the global vertical axis projected to the sagittal plane of the shank's LCS (Fig. 3) as follows:

$$\theta_{shank} = cos^{-1}\left({e_{shank,z}}^{T} e_{ver,sag}\right), \qquad (1)$$

where $\boldsymbol{e}_{ver,sag}$ is the global vertical axis projected to the sagittal plane of the LCS of the shank (Fig. 3). A positive value indicates that the shank segment inclined anteriorly relative to the global vertical axis in the sagittal plane of the LCS of the shank.

The GRF inclination angle $\theta_f$ was defined as follows:

$$\theta_f = cos^{-1}\left({f_{sag}}^{T} e_{ver,sag} / |f_{sag}|\right), \qquad (2)$$

where $\boldsymbol{f}_{sag}$ is the GRF vector projected on the sagittal plane of the LCS of the shank (Fig. 3).

The CoP anterior/posterior position relative to the shank proximal/distal axis was defined as follows:

$$r_{cop,x} = {e_{shank,x}}^{T} r_{cop}, \qquad (3)$$

where $\boldsymbol{r}_{cop}$ is the position vector from KJC to the CoP. When $r_{cop,x}$ > 0, the CoP is anterior to the shank proximal/distal axis and vice versa. The knee resultant moment vector $\boldsymbol{\tau}$ was calculated using the Newton-Euler equation of motion; then, the knee resultant flexion (+)/ extension (–) moment was defined as follows:

$$\tau_y = {e_{knee,y}}^T \tau, \quad (4)$$

The knee flexion(+)/extension(–) moment due to GRF $\boldsymbol{\tau}_{f,y}$ was defined as follows:

$$\tau_{f,y} = {e_{knee,y}}^T \left(r_{cop} \times f\right), \quad (5)$$

where $\boldsymbol{f}$ is GRF vector and (×) denotes operator of cross product.

The anterior/posterior component of the GRF vector $f_{a/p}$ relative to the shank proximal/distal axis was defined as a projection of the GRF vector $\boldsymbol{f}$ onto the shank anterior/posterior axis $\boldsymbol{e}_{shank,x}$ as follows:

$$f_{\frac{a}{p}} = {e_{shank,x}}^T f, \quad (6)$$
